# Evaluating the Performance of ChatGPT in Ophthalmology: An Analysis of its Successes and Shortcomings

**DOI:** 10.1101/2023.01.22.23284882

**Authors:** Fares Antaki, Samir Touma, Daniel Milad, Jonathan El-Khoury, Renaud Duval

## Abstract

We tested the accuracy of ChatGPT, a large language model (LLM), in the ophthalmology question-answering space using two popular multiple choice question banks used for the high-stakes Ophthalmic Knowledge Assessment Program (OKAP) exam. The testing sets were of easy-to-moderate difficulty and were diversified, including recall, interpretation, practical and clinical decision-making problems. ChatGPT achieved 55.8% and 42.7% accuracy in the two 260-question simulated exams. Its performance varied across subspecialties, with the best results in general medicine and the worst in neuro-ophthalmology and ophthalmic pathology and intraocular tumors. These results are encouraging but suggest that specialising LLMs through domain-specific pre-training may be necessary to improve their performance in ophthalmic subspecialties.

## INTRODUCTION

Since 2015, significant progress has been made in the application of artificial intelligence (AI) and deep learning (DL) in medicine, particularly in ophthalmology^1^. DL has been widely used for image recognition using various types of ophthalmic data, such as fundus photographs and optical coherence tomography, and has shown strong results in detecting a wide range of diseases^2 3^. More recently, there has been growing interest in using DL for natural language processing (NLP) in ophthalmology, which involves using AI to understand and interact with human language^4^.

NLP has received considerable media attention in the past months due to the release of large DL models called foundation models^5^. These models can be repurposed for various tasks, such as generating text or images, after being trained on a wide range of unlabelled data^6 7^. A prominent example of a foundation model is Generative Pre-trained Transformer 3 (GPT-3), a large language model (LLM) that generates human-like text. It is based on the transformer architecture, and was trained on a massive dataset of text (>400 billion words) from the internet including books, articles, and websites^8^.

There has been recent interest in evaluating the capabilities of LLMs for understanding and generating natural language in medicine^9 10^. The medical domain can pose a significant challenge for LLMs since clinical reasoning often requires years of training and hands-on experience to master. In 2022, Singhal and colleagues demonstrated the capabilities of PaLM, a 540-billion parameter LLM, by testing it on multiple-choice questions from the US Medical Licensing Exam (USMLE) with an impressive 67.6% accuracy^9^. More recently, Kung and colleagues evaluated the performance of ChatGPT, a generic LLM developed by OpenAI that is based on the GPT-3 series and optimised for dialogue, using multiple-choice questions also from USMLE^11^. They found that ChatGPT achieved overall accuracy above 50% in most of their experiments, and also provided insightful explanations to support its answer choices.

To our knowledge, the performance of LLMs has not yet been examined in the ophthalmology question-answering space. In this study, we evaluated the performance of ChatGPT in ophthalmology by using two popular board preparation question banks: the American Academy of Ophthalmology’s Basic and Clinical Science Course (BCSC) Self-Assessment Program and the OphthoQuestions online question bank. These resources have been shown to be effective in studying for board exams and have been linked to improved performance on the standardised Ophthalmic Knowledge Assessment Program (OKAP) exam, which is taken annually by ophthalmology residents in the United States and Canada^12 13^.

## METHODS

### ChatGPT

We used the free research preview of ChatGPT January 9 version (OpenAI, San Francisco). ChatGPT is a fine-tuned LLM based on a model from the GPT-3.5 series^14^. GPT-3 has a transformer architecture and was trained using billions of text data obtained from writings on the internet. This process is done by training the model to minimise the difference between the predicted word and the actual word in the training dataset. Once the model is trained, it can be used to generate new text by providing it with a prompt and allowing it to predict the next word. The model then uses this predicted word as the context for the next prediction, and this process is repeated until a complete sentence or paragraph is generated^8^. ChatGPT goes beyond just predicting the next word, as it is optimised for dialogue and was trained using human feedback. This allows it to understand and respond to human expectations when answering questions^15^.

### BCSC and OphthoQuestions

In January 2023, we generated a test set of 260 questions from the BCSC Self-Assessment Program and 260 questions from OphthoQuestions through personal subscription accounts. Those questions are not publicly accessible thereby excluding the possibility of prior indexing in any search engine (like Google) or in the ChatGPT training dataset. For the BCSC and OphthoQuestions test sets, we randomly generated 260 questions out of a pool of 4,458 and 4,539 potential questions, respectively. During the process, any questions that included visual information such as clinical, radiologic, or graphical images were removed and replaced since ChatGPT does not currently support such data. We generated 20 random questions from each of the 13 sections of the OKAP exam: Update on General Medicine, Fundamentals and Principles of Ophthalmology, Clinical Optics and Vision Rehabilitation, Ophthalmic Pathology and Intraocular Tumors, Neuro-Ophthalmology, Pediatric Ophthalmology and Strabismus, Oculofacial Plastic and Orbital Surgery, External Disease and Cornea, Uveitis and Ocular Inflammation, Glaucoma, Lens and Cataract, Retina and Vitreous, and Refractive Surgery.

### Question format and encoding

We aimed to replicate an OKAP exam, and therefore maintained the standard multiple-choice format with one correct answer and three incorrect options (distractors). We employed a zero-shot approach for the lead-in prompt, using the prompt “Please select the correct answer and provide an explanation” followed by the question and answer options, without providing any examples^9^. Although more challenging for ChatGPT^8^, we chose this technique as it is the closest to human test-taking. A new session was started in ChatGPT for each question to reduce memory retention bias.

### Level of cognition and question difficulty

Since the BCSC and OphthoQuestions questions were not labelled for difficulty, we labelled them according to the cognitive level and calculated a difficulty index^16^. We did this to analyse ChatGPT’s performance based on not only the subject, but also the type of question and level of difficulty. Despite having no control over the distribution of cognitive level and question difficulty in each of the randomly-generated test sets, we elected not to balance them manually to prevent cherry-picking, thereby avoiding bias in the experiment results.

We used a simplified scoring system of Low and High cognitive level, instead of the three-tier system proposed in the OKAP User’s Guide^17^. This was done because we found it difficult to distinguish between Level 2 and Level 3 questions, and we wanted to avoid making assumptions about the intended goal of the questions. Low cognitive level questions tested recall of facts and concepts, such as identifying the gene implicated in a known condition. High cognitive level questions tested the ability to interpret data, make calculations and manage patients, like in common clinical optics exercises (e.g. cylinder transpositions) or to select the best treatment for specific cancers in unique clinical contexts (e.g. the optimal treatment for metastatic sebaceous cell carcinoma of the eyelid). The difficulty index represented the percentage of individuals who correctly answered a question, as reported by BCSC and OphthoQuestions platforms for each question. Questions with a higher difficulty index are considered easier. The questions were categorized into three levels of difficulty: difficult (<30%), moderate (≥ 30% and <70%), and easy (≥ 70%)^18^.

### Statistical analysis

Accuracy was determined by comparing ChatGPT’s answer to the answer key provided by the question banks. We used logistic regression (all the input variables were entered simultaneously) to examine the effect of the exam section, cognitive level, and difficulty index on ChatGPT’s answer accuracy. We then performed a post hoc analysis using Tukey’s test to determine if there were significant differences in accuracy between exam sections while controlling for questions difficulty and cognitive level. By controlling for those factors, we were able to isolate the effect of the exam section on accuracy and determine if there were any meaningful differences between the tested topics.

## RESULTS

### The testing sets demonstrated similar difficulty and cognitive levels

The BCSC and OphthoQuestions training sets had a similar level of difficulty (p=0.154) and mostly included easy and moderate questions in a very similar distribution (p=0.102), as illustrated in **Figure 1** and **Table 1**. Likewise, the questions’ cognitive levels were comparable between the two test sets (p=0.425). Those similarities allowed us to combine the testing sets during further analyses.

**Table 1:**
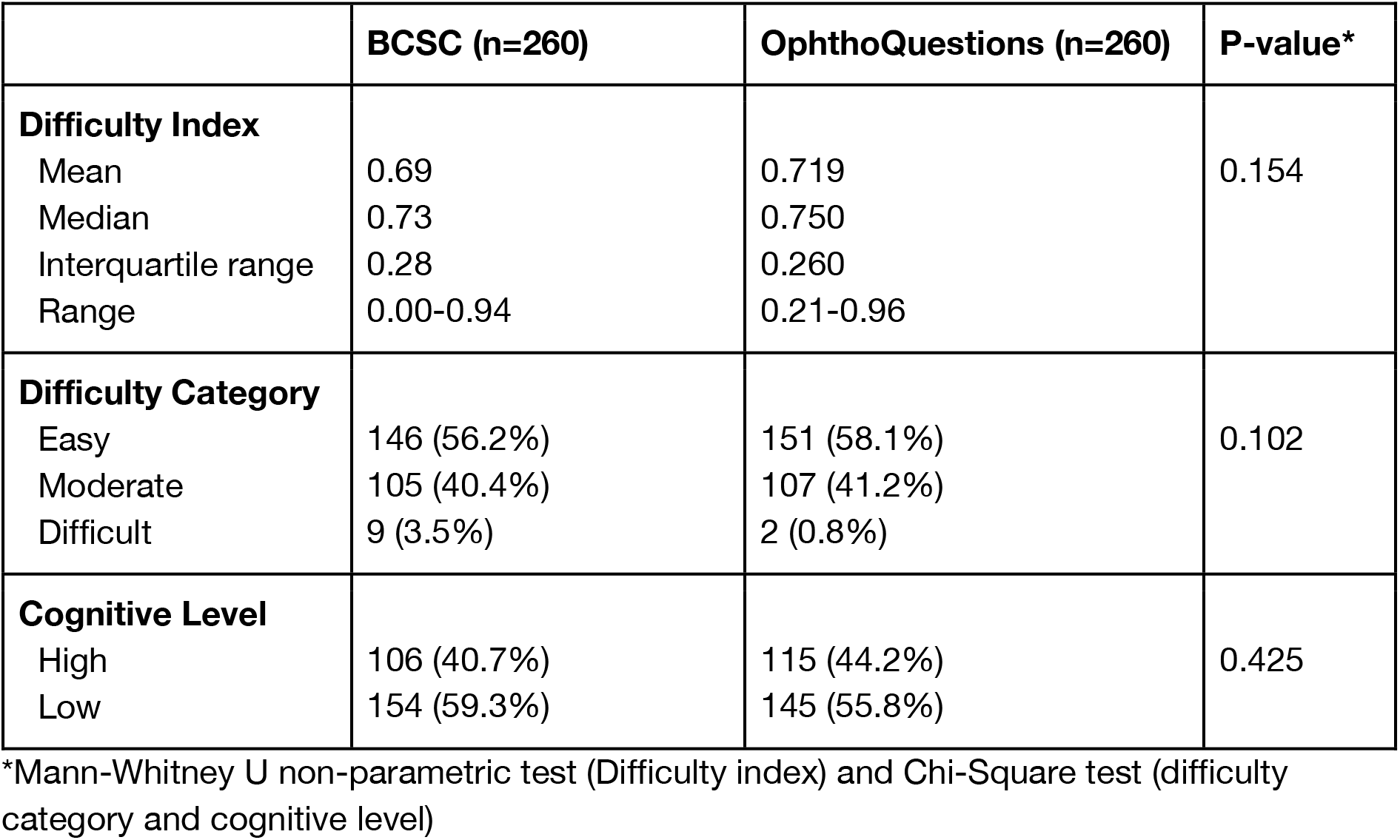
Baseline characteristics of the testing sets.

**Figure 1:**
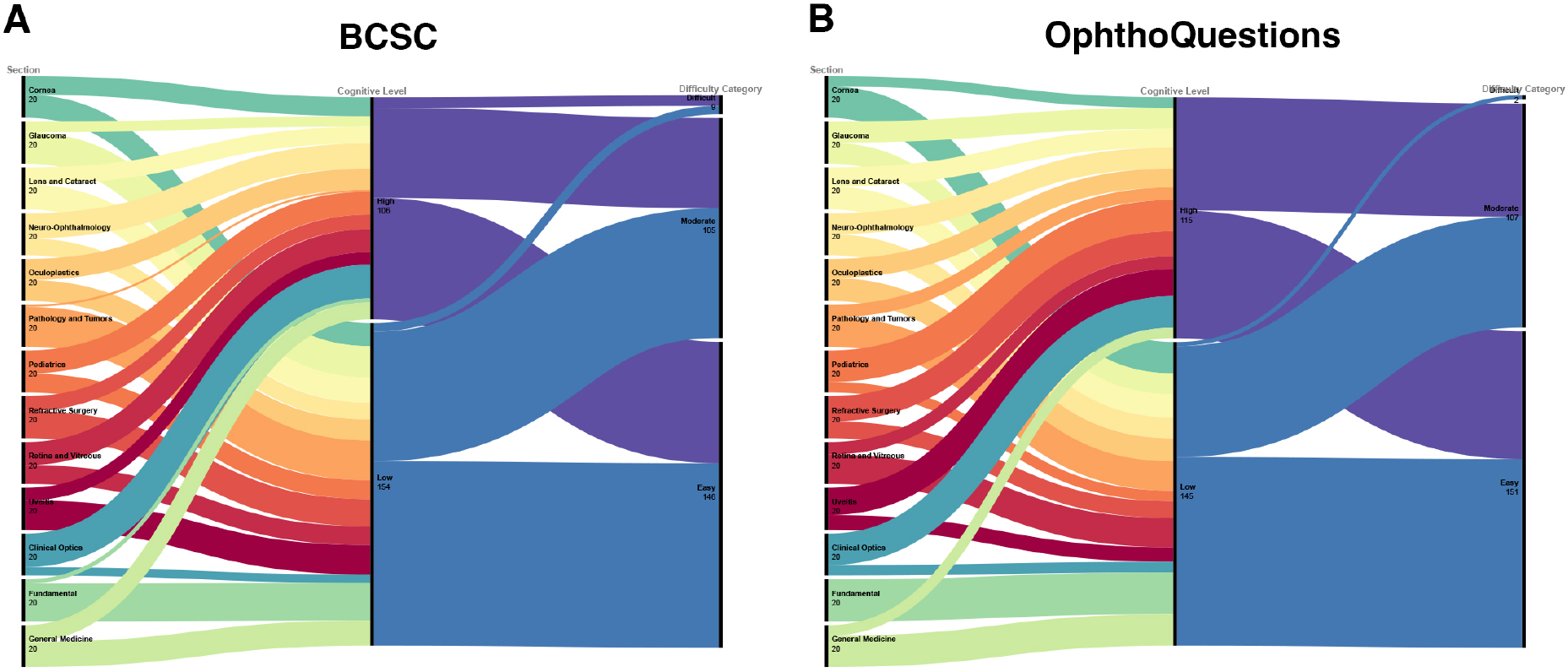
Alluvial diagram illustrating the distribution of questions across exam sections, cognitive level, and question difficulty. Despite having been generated at random, the BCSC and OphthoQuestions test sets have a similar distribution of questions with High and Low cognitive levels and similar difficulty.

### ChatGPT had a modest overall performance with variability between testing sets

ChatGPT had an accuracy of 55.8% on the BCSC set and 42.7% on the OphthoQuestions set. As shown in **Figure 2**, it performed well in General Medicine (75%), Fundamentals (60%) and Cornea (60%), but not as well in Neuro-ophthalmology (25%), Glaucoma (37.5%), and Pediatrics and Strabismus (42.5%). Although there were variations in performance between the BCSC and OphthoQuestions for the same subject, the difference was only statistically significant for Pediatric Ophthalmology & Strabismus (p=0.010) (**Supplemental Table 1**).

**Figure 2:**
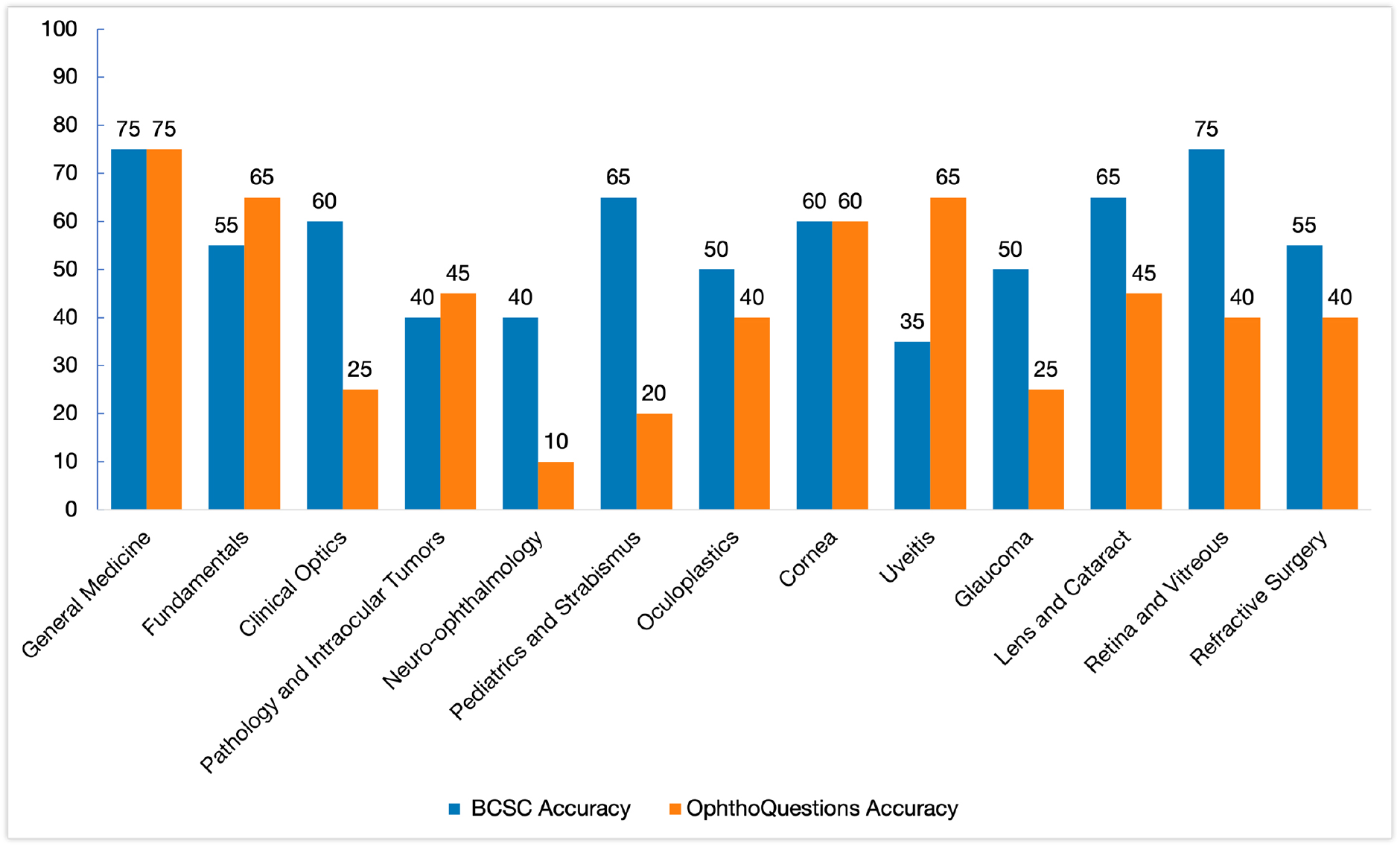
Bar plot of the accuracy of ChatGPT across exam sections for the BCSC and OphthoQuestions testing sets. Despite the variations in performance between the testing sets for the same subject, the difference was only statistically significant for Pediatric Ophthalmology & Strabismus (p=0.010).

### ChatGPT’s accuracy depends on the exam section and question difficulty

Our logistic regression analysis showed that the exam section (LR 27.57, p=0.006) followed by question difficulty (LR 24.05, p<0.001) were most predictive of ChatGPT’s answers accuracy (**Table 2**). **Supplemental Table 2** provides the results of the analysis per testing set. While controlling for question difficulty and cognitive level, we found significant differences in ChatGPT’s performance between General Medicine and each of Glaucoma (p=0.002), Neuro-Ophthalmology (p<0.001), and Ophthalmic Pathology and Intraocular Tumors (p=0.029) (**Supplemental Figure 1**). Similarly, we found that accuracy improved with increased difficulty index (easier questions) even when controlling for the exam section and cognitive level. **Supplemental Figures 2 and 3** provides the results of the post hoc analysis for each of the testing sets.

**Table 2:** Likelihood ratio test for exam section, cognitive level, and difficulty index for all questions.

| Effects | LR Chisq | Df | Pr(>Chisq) |
| --- | --- | --- | --- |
| Section | 27.57 | 12 | 0.006* |
| Cognitive Level | 3.54 | 1 | 0.06 |
| Difficulty index | 24.05 | 1 | <0.001* |

\* Statistically significant at the 0.05 level

## DISCUSSION

In the past months, there has been significant interest in examining the utility of LLMs in medicine^5^. Despite having encouraging impacts in various industries, it is important to thoroughly evaluate their performance and biases before determining their clinical usefulness^4 19^. In this study, we provide evidence on the performance of ChatGPT, a non-domain specific LLM, in responding to questions found on the OKAP exam.

ChatGPT achieved an accuracy of 55.8% on the simulated OKAP exam using the BCSC testing set and 42.7% on the OphthoQuestions testing set. On average, humans score 74% on the BCSC question bank and 61% on OphthoQuestions, with first-year residents scoring an average of 53% on OphthoQuestions. ChatGPT is therefore within range to perform at the level of an average first-year resident. We believe this outcome is noteworthy and promising within ophthalmology, as our results approach ChatGPT’s performance on the USMLE despite being a much more specialised examination^11^. Our findings are also encouraging as ChatGPT’s accuracy in ophthalmology is similar to the typical accuracy seen in general medical question answering by state-of-the-art LLMs, typically around 40-50% as reported in publications from 2022^9^.

We found that ChatGPT’s accuracy mostly depended on the exam section, even when controlling for question difficulty and cognitive level. The highest accuracy was in General Medicine (75%) and the second highest was in Fundamentals (60%). The model’s high performance in these areas might be attributed to the vast amount of training data and resources available on the internet for those topics. In contrast, ChatGPT performed poorest in Neuro-ophthalmology and Ophthalmic Pathology and Intraocular Tumors. Those are highly-specialized domains that are considered challenging even within the ophthalmology community. For example, up to 40% of patients referred to a neuro-ophthalmology subspecialty service are misdiagnosed^20^, and similar referral patterns are observed in ocular oncology^21^.

Understanding why ChatGPT makes mistakes is important. We found that question difficulty was predictive of ChatGPT’s accuracy, even when controlling for the exam section and cognitive level. ChatGPT was more accurate when a higher percentage of human peers obtained the right answer for a specific question. This discovery is comforting as it suggests that ChatGPT’s responses align, to a certain degree, with the collective understanding of ophthalmology trainees. In parallel, Kung and colleagues showed that the accuracy of ChatGPT is heavily influenced by concordance and insight, indicating that inaccurate responses are caused by a lack of training information for the USMLE^11^. We plan to perform a similar qualitative analysis to identify areas for improvement in the ophthalmology space. Incorporating ChatGPT with other specialised foundation models that are trained using domain-specific sources (like EyeWiki) might be required to improve its accuracy.

Despite its encouraging performance, the imminent implementation of ChatGPT in ophthalmology may be limited because it does not have the capability to process images. This is a significant limitation as ophthalmology is a field that heavily relies on visual examination and imaging to diagnose, treat, and monitor patients. LLMs like ChatGPT may need to incorporate other transformer models that can handle multiple types of data, such as the Contrastive Language-Image Pretraining (CLIP) model^22^, which can classify images and generate a text description that ChatGPT can then use to respond to a question. While this approach shows potential, it is limited by its reliance on a large amount of image-text pairs from the internet (in the case of CLIP) that are not specific to our domain. These data may not be sufficient to accurately distinguish subtle and specific differences relevant to medicine and ophthalmology^23^. For instance, CLIP may not be able to accurately caption a “superior” retinal detachment that would need a pneumatic retinopexy, as opposed to an “inferior” retinal detachment that might require a scleral buckle.

As the performance of ChatGPT improves (perhaps through prompting strategies), it will be important to work collectively toward building safeguards for our patients^4^. Those will include protecting vulnerable populations from biases and evaluating the potential harm or risk of acting on the answers provided by LLMs like ChatGPT. This will be particularly important for high-level decision-making questions that may be challenging to train for due to inconclusive training data on the internet, reflecting the variability in research data as well as global practice patterns. We are excited about the potential of ChatGPT in ophthalmology, but we remain cautious when considering the potential clinical applications of this technology.

## Data Availability

All data produced in the present study are available upon reasonable request to the authors. The specific question sets are proprietary of the BCSC Self-Assessment Program and OphthoQuestions.

## ACKNOWLEDGEMENTS

We express our gratitude to Mr. Charles-Édouard Giguère, statistician at the Institut Universitaire en Santé Mentale de Montréal, for his assistance in the statistical analysis.

## AUTHOR CONTRIBUTIONS

FA, ST, DM and RD conceptualised the study and designed the experiments. FA, ST, DM and JE collected the data. FA analysed the data. FA and JE drafted the initial manuscript. FA designed the tables and figures. All authors reviewed and discussed the results. All authors edited and revised the manuscript before approving the final version of this manuscript.

