## Supplemental Table 1 for "Evaluating the Performance of ChatGPT in Ophthalmology: An Analysis of its Successes and Shortcomings"

**Supplemental Table 1: ChatGPT's performance per exam section of each of the BCSC and OphthoQuestions testing sets**

| | BCSC (n=260) | OphthoQuestions (n=260) | P value* |
|---|---|---|---|
| **General Medicine** | | | 1.000 |
| Correct | 15 (75%) | 15 (75%) | |
| Incorrect | 5 (25%) | 5 (25%) | |
| **Fundamentals** | | | 0.748 |
| Correct | 11 (55%) | 13 (65%) | |
| Incorrect | 9 (45%) | 7 (35%) | |
| **Clinical Optics** | | | 0.054 |
| Correct | 12 (60%) | 5 (25%) | |
| Incorrect | 8 (40%) | 15 (75%) | |
| **Pathology and Intraocular Tumors** | | | 1.000 |
| Correct | 8 (40%) | 9 (45%) | |
| Incorrect | 12 (60%) | 11 (55%) | |
| **Neuro-ophthalmology** | | | 0.065 |
| Correct | 8 (40%) | 2 (10%) | |
| Incorrect | 12 (60%) | 18 (90%) | |
| **Pediatrics and Strabismus** | | | 0.010** |
| Correct | 13 (65%) | 4 (20%) | |
| Incorrect | 7 (35%) | 16 (80%) | |
| **Oculoplastics** | | | 0.751 |
| Correct | 10 (50%) | 8 (40%) | |
| Incorrect | 10 (50%) | 12 (60%) | |
| **Cornea** | | | 1.000 |
| Correct | 12 (60%) | 12 (60%) | |
| Incorrect | 8 (40%) | 8 (40%) | |
| **Uveitis** | | | 0.113 |
| Correct | 7 (35%) | 13 (65%) | |
| Incorrect | 13 (65%) | 7 (35%) | |

| | | | |
|---|---|---|---|
| **Glaucoma**<br>Correct<br>Incorrect | <br>10 (50%)<br>10 (50%) | <br>5 (25%)<br>15 (75%) | 0.191 |
| **Lens and Cataract**<br>Correct<br>Incorrect | <br>13 (65%)<br>7 (35%) | <br>9 (45%)<br>11 (55%) | 0.341 |
| **Retina and Vitreous**<br>Correct<br>Incorrect | <br>15 (75%)<br>5 (25%) | <br>8 (40%)<br>12 (60%) | 0.054 |
| **Refractive Surgery**<br>Correct<br>Incorrect | <br>11 (55%)<br>9 (45%) | <br>8 (40%)<br>12 (60%) | 0.527 |

*Fisher's Exact test

**Statistically significant at the 0.05 level
