## Supplemental Table 2 for "Evaluating the Performance of ChatGPT in Ophthalmology: An Analysis of its Successes and Shortcomings"

**Supplemental Table 2: Likelihood ratio test for exam section, cognitive level, and difficulty index for each of the BCSC and OpthoQuestions sets**

| **Effects** | **LR Chisq** | **Df** | **Pr(>Chisq)** |
|---|---|---|---|
| **BCSC** | | | |
| Section | 22.87 | 12 | 0.029* |
| Cognitive level | 8.17 | 1 | 0.004* |
| Difficulty index | 14.70 | 1 | <0.000* |
| **OphthoQuestions** | | | |
| Section | 35.46 | 12 | <0.001* |
| Cognitive level | 0.001 | 1 | 0.98 |
| Difficulty index | 13.38 | 1 | <0.001* |

* Statistically significant at the 0.05 level
