## Supplemental Figure 1 for "Evaluating the Performance of ChatGPT in Ophthalmology: An Analysis of its Successes and Shortcomings"

**Supplemental Figure 1: Post hoc analysis using Tukey's test to isolate the effect of exam section and difficulty index using all questions. (A)** Bar plot of the percentage of accuracy by exam section. The blue square brackets identify the significant differences. The significant contrasts were the following: General Medicine – Glaucoma (estimate 0.424, p=0.002), General Medicine - Neuro-Ophthalmology (estimate 0.486, p<0.001) and General Medicine - Pathology and Tumors (estimate 0.367, p=0.029). **(B)** Predicted percentage of accuracy by difficulty index. Accuracy increased with increasing difficulty index (easier questions).

**(A)**

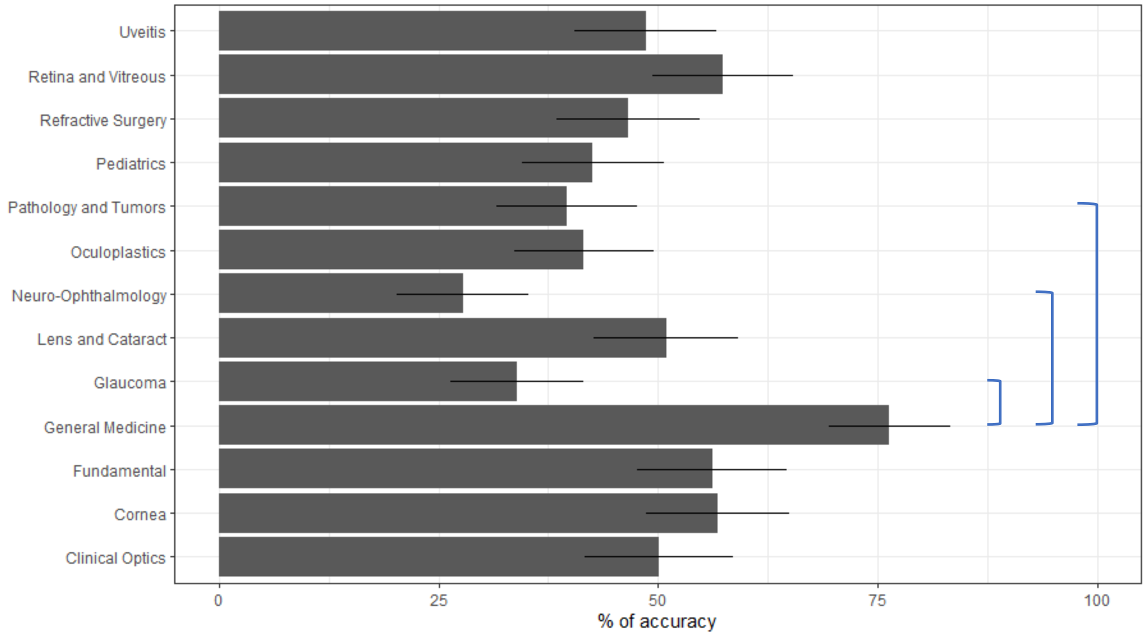


**(B)**

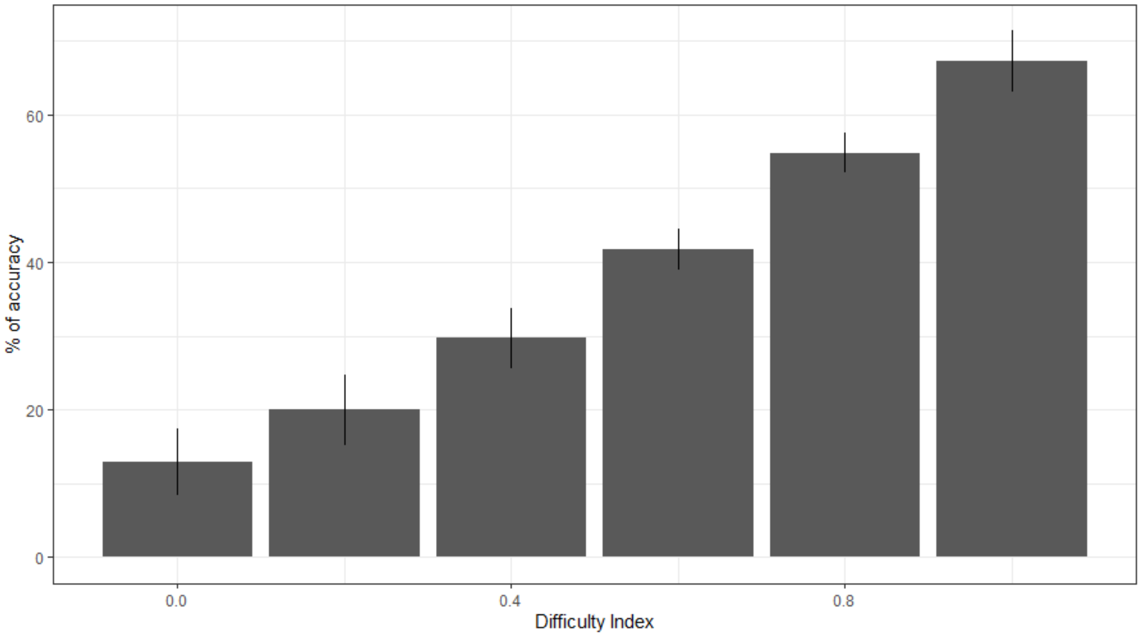
