## Supplemental Figure 2 for "Evaluating the Performance of ChatGPT in Ophthalmology: An Analysis of its Successes and Shortcomings"

**Supplemental Figure 2: Post hoc analysis using Tukey's test to isolate the effect of exam section, cognitive level and difficulty index using the BCSC questions. (A)** Bar plot of the percentage of accuracy by exam section. The blue square brackets identify the significant differences. The significant contrasts were the following: Clinical Optics – Uveitis (estimate 0.473, p = 0.042), General Medicine - Pathology and Tumors (estimate 0.487, p=0.029) and General Medicine – Uveitis (estimate 0.508, p = 0.013). **(B)** Predicted percentage of accuracy by difficulty index. Accuracy increased with increasing difficulty index (easier questions). **(C)** Predicted percentage of accuracy comparing Low and High cognitive levels. Higher accuracy was associated to Low cognitive level questions.

**(A)**

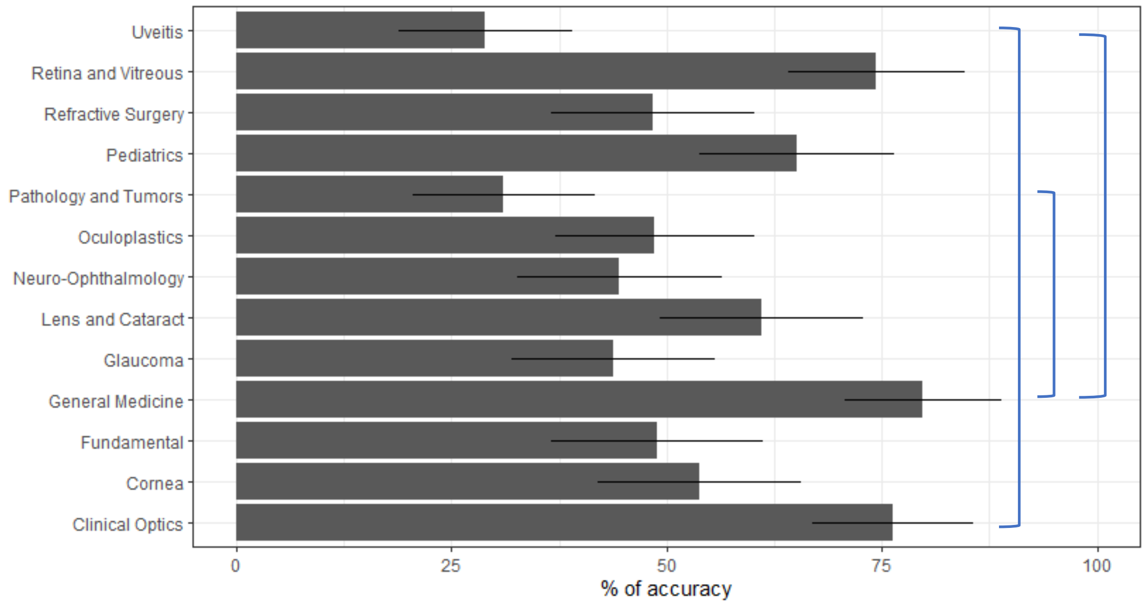


**(B)**

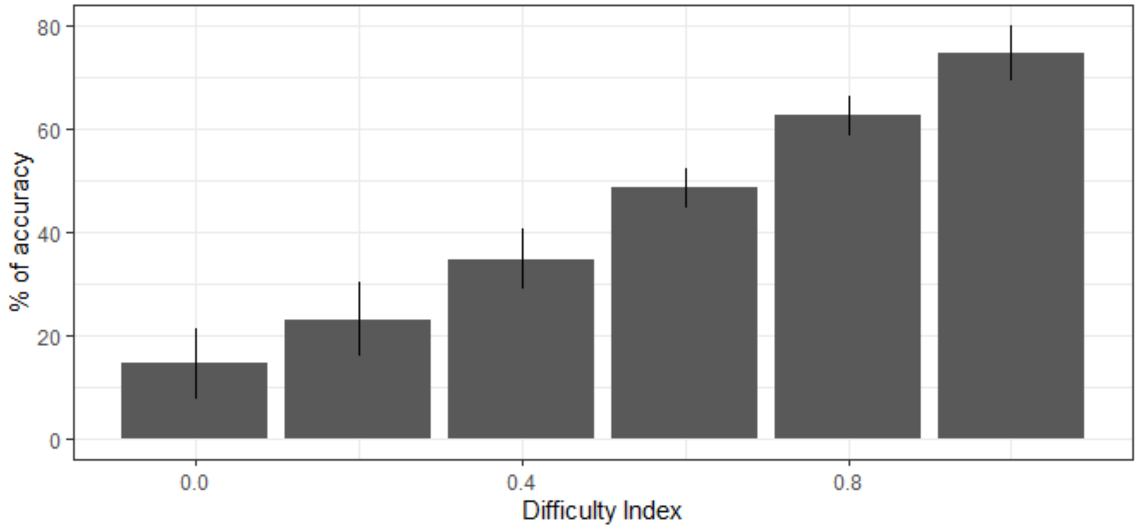

(C)

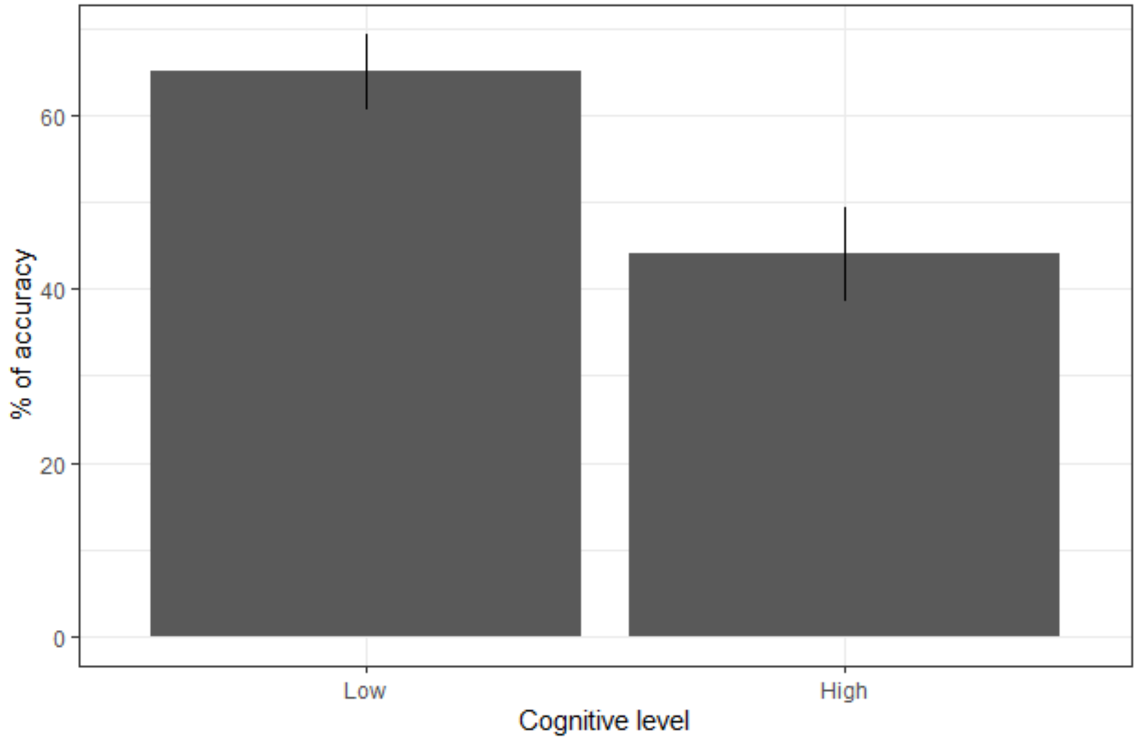
