## Supplemental Figure 3 for "Evaluating the Performance of ChatGPT in Ophthalmology: An Analysis of its Successes and Shortcomings"

**Supplemental Figure 3: Post hoc analysis using Tukey's test to isolate the effect of exam section and difficulty index using the OphthoQuestions questions. (A)** Bar plot of the percentage of accuracy by exam section. The blue square brackets identify the significant differences. The significant contrasts were the following: Clinical Optics - General Medicine (estimate -0.501, p=0.028), Cornea - Neuro-Ophthalmology (estimate 0.506, p=0.013), Fundamentals - Neuro-Ophthalmology (estimate 0.518, p=0.016), General Medicine - Glaucoma (estimate 0.538, p=0.004), General Medicine - Neuro-Ophthalmology (estimate 0.638, p=<0.001), General Medicine - Pediatrics (estimate 0.552, p=0.004) and Neuro-Ophthalmology- Uveitis (estimate -0.536, p=0.004). **(B)** Predicted percentage of accuracy by difficulty index. Accuracy increased with increasing difficulty index (easier questions).

**(A)**

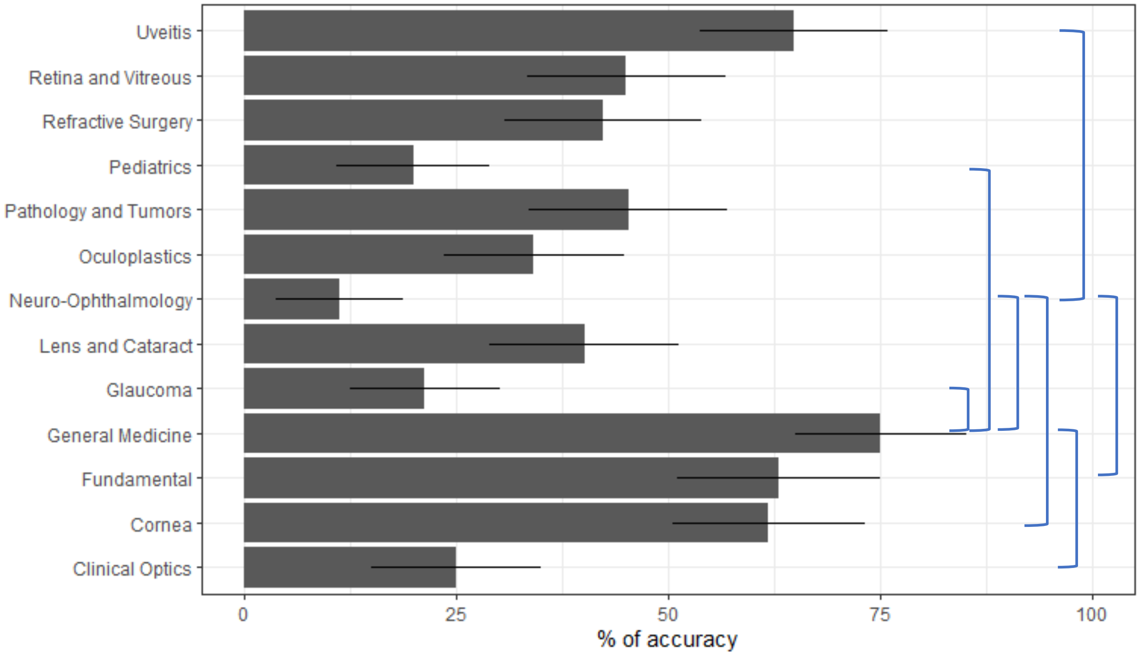


**(B)**

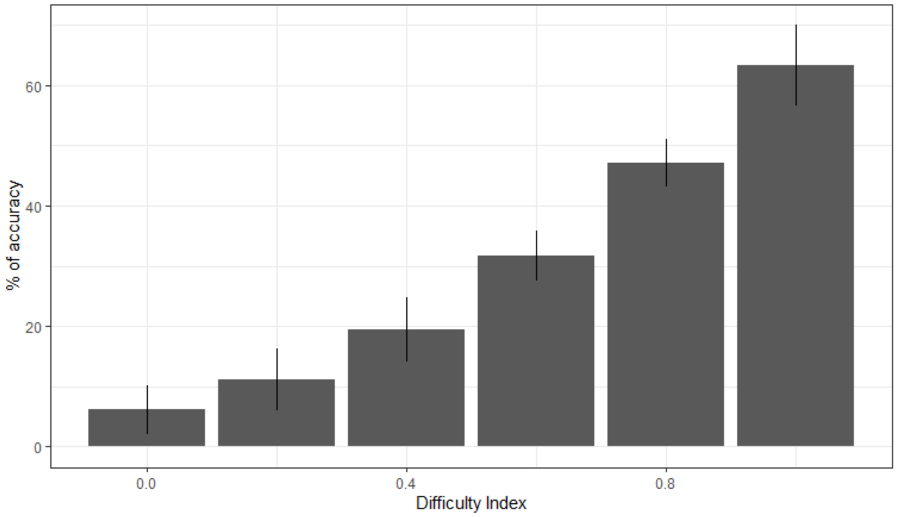
